# Wastewater-based Epidemiology for Averting COVID-19 Outbreaks on The University of Arizona Campus

**DOI:** 10.1101/2020.11.13.20231340

**Authors:** Walter W. Betancourt, Bradley W. Schmitz, Gabriel K. Innes, Kristen M. Pogreba Brown, Sarah M. Prasek, Erika R. Stark, Aidan R. Foster, Ryan S. Sprissler, David T. Harris, Samendra P. Sherchan, Charles P. Gerba, Ian L. Pepper

## Abstract

The University of Arizona utilized wastewater-based epidemiology paired with clinical testing as a surveillance strategy to monitor COVID-19 prevalence in a dormitory community. Positive SARS-CoV-2 RNA detection in wastewater led to prompt testing of all residents and the identification and isolation of three infected individuals which averted potential disease transmission.

**Text:** Wastewater-based epidemiology (WBE) utilizes concentrations of SARS-CoV-2 in sewage to monitor population-level COVID-19 infections *(1–3)*. Currently, WBE is a promising indicator to support public health decisions *(3,4)*. In this case study, WBE was used to detect a COVID-19 outbreak in a student dormitory (henceforth Dorm A) at the University of Arizona (UArizona).

**The Study:** UArizona incorporated wastewater surveillance as a potential early-warning tool for COVID-19 outbreaks on campus. Grab samples (1L) were collected from a sewer manhole specific to Dorm A, between August 18-31 to monitor SARS-CoV-2 RNA in wastewater. Upon positive detection of viral RNA in wastewater samples, clinical testing was conducted on every individual living in the dorm. UArizona performed two clinical testing modalities, antigen (1 hour turnaround) test via anterior nasal swab and RT-PCR (48-72 hour turnaround) via nasopharyngeal swab samples. Individuals were subject to clinical testing via two routes: Campus Health Services (CHS) if experiencing symptoms or Test All Test Smart (TATS) regardless of symptoms. Refer to Appendix for method details.

**Article Summary Line:** Wastewater-based epidemiology with subsequent clinical testing identified individuals infected with COVID-19 living in a dormitory and further spread of disease was prevented with public health action.

---

On March 13, 2020, UArizona advised students and employees to work remotely. During the summer, UArizona administration assembled a Task Force and Campus Re-Entry Working Groups to prepare for students’ safe return. Seven expert teams were created consisting of COVID-19 clinical testing, thermometry and wellness checks, contact tracing, healthcare and guidance, isolation, data platforms and communication, and WBE. Utilization of WBE in conjunction with clinical testing was seen to be critical for the early detection of infections in student dorms.

The WBE expert team hypothesized that surveillance of defined communities in dorms would provide an effective means for identifying new cases of COVID-19 since 1) each dorm contained a known population, 2) dorm students provide a representation of the overall status of campus health, 3) wastewater samples could be collected from individual buildings, and 4) actionable public health responses could be initiated in the event of positive wastewater detection. Wastewater samples were collected from sewer manholes downstream from each dorm prior to convergence or mixing with other sewer lines, resulting in samples specific to individual buildings with defined communities.

On August 17, students were permitted to move into Dorm A (Figure 1). Before entering on-campus housing, students were required to test negative via UArizona’s COVID-19 nasal swab antigen test and follow public health guidelines, such as wearing a mask and committing to social distancing. Students testing positive were prohibited from entering the dorm and were required to remain in isolation until remaining symptom-free for a minimum of 10 days, per Centers for Disease Control and Prevention (CDC) guidelines *(5)*.

**Figure 1.**
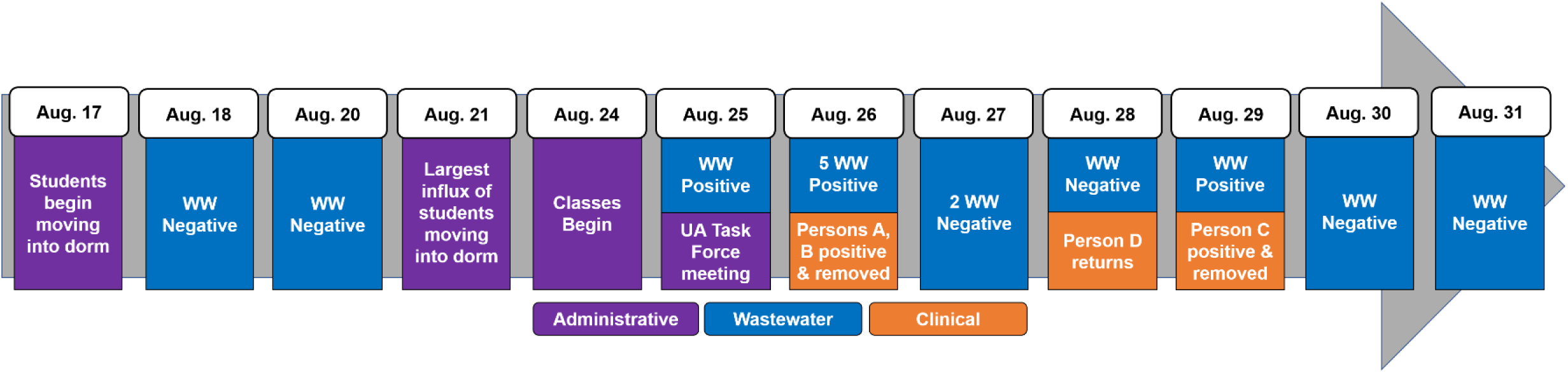
Timeline of events at Dorm A. Dates (left to right) and events (top to bottom) are listed in chronological order. WW = wastewater.

A baseline survey of SARS-CoV-2 RNA in wastewater was conducted prior to the start of the fall semester by collecting wastewater samples from Dorm A on August 18 and 20. No SARS-CoV-2 RNA was detected in these samples. Negative results corresponded with the requirement that all students test negative prior to entering the dorm. The largest number of students that moved into campus housing occurred on August 21, three days before the start of classes on August 24 (Figure 1). On August 25, wastewater from Dorm A tested positive for SARS-CoV-2 N1 gene (1.61 x 105 copies/L) signaling the presence of the virus in the dorm (Table 1). This positive sample triggered an emergency UA Task Force meeting, which supported additional wastewater sampling and clinical testing among Dorm A residents (Figure 1).

**Table 1.**
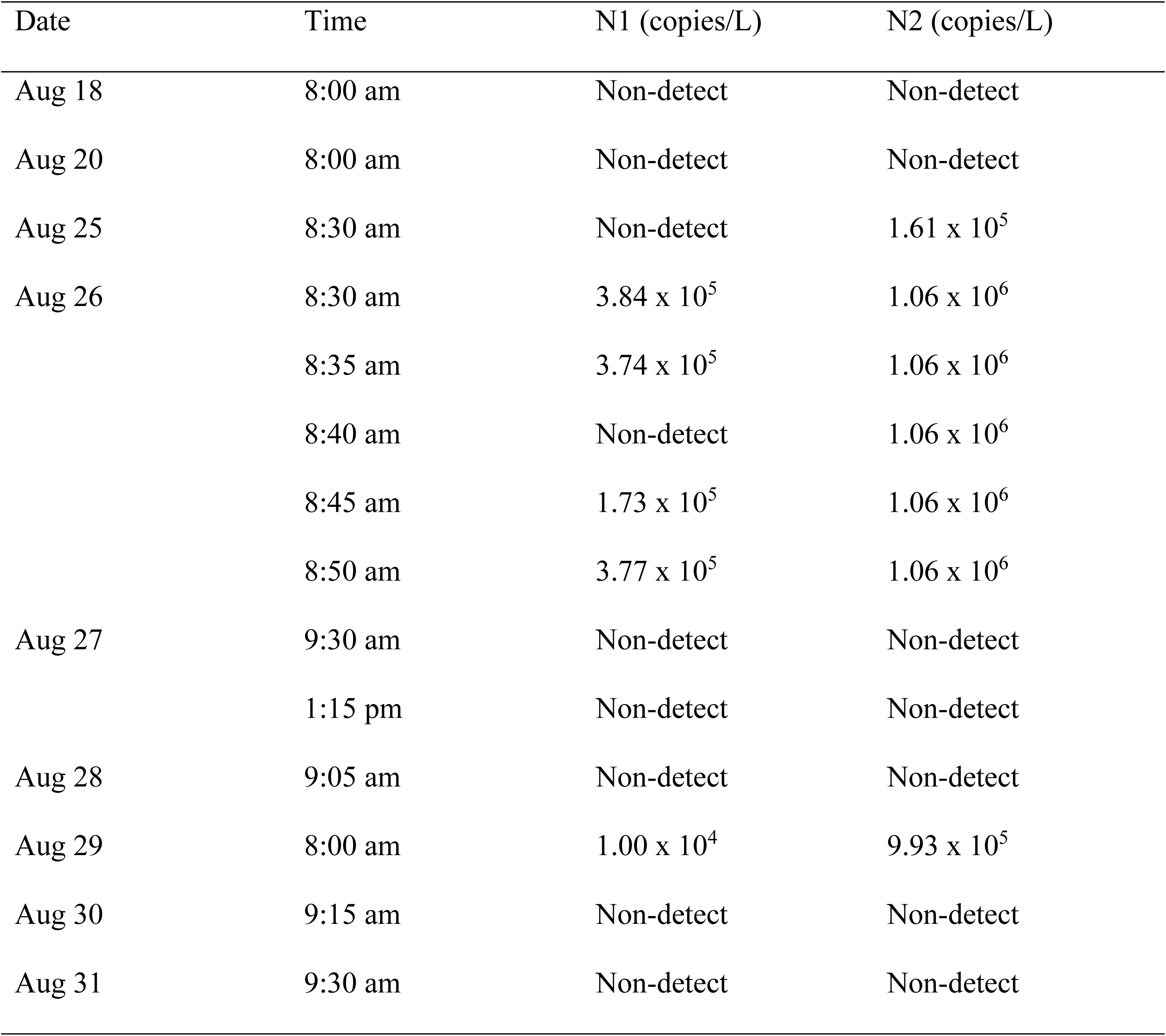
Wastewater surveillance from manhole samples at Dorm A.

The next day (August 26), five wastewater samples were collected once every five minutes between 8:30-8:50 am. Importantly, all five samples yielded virtually identical SARS-CoV-2 concentrations (Table 1). This validated homogenous virus dispersion within the sewer environment *(2,6)*, which suggested that grab samples were representative, and therefore composite sampling was unnecessary. That same day, 270 antigen tests and 260 PCR tests were conducted on-site at the dorm (via TATS) out of 311 total residents (Table 2). Antigen testing identified one positive individual (Person A) despite the individual showing no symptoms. The other 269 antigen tests were negative. Simultaneously, another individual (Person B) reported to CHS experiencing symptoms and tested positive via an antigen test. Person A and Person B were immediately relocated into an isolation facility to prevent transmission of the virus. Another PCR test was inconclusive (Person C) and retested per CDC guidelines *(7)*. Results of this retest were not available until August 29.

**Table 2.** COVID-19 clinical testing results for persons living in Dorm A.

| Date | CHS Antigen | CHS PCR | TATS Antigen | TATS PCR | Total<br>Positive |
| --- | --- | --- | --- | --- | --- |
| Aug 24 | - | 0/1 | - | - | 0 |
| Aug 25 | 0/1 | - | 0/2 | - | 0 |
| Aug 26 | 1/4 | 1/4 | 1/270 | 2/260 | 3 |
| Aug. 27 | 0/1 | 0/1 | 0/26 | 0/21 | 0 |
| Aug 28 | 0/2 | 0/2 | 0/5 | - | 0 |
| Aug 29 | - | - | - | - | 0 |
| Aug 30 | - | - | - | - | 0 |
| Aug 31 | 0/2 | 0/1 | - | - | 0 |

No clinical tests were performed at the dorm prior to August 24, since all individuals tested negative at a designated testing site on-campus prior to entering the dormitory between August 17 and 23. CHS = campus health service; TATS = Test All Test Smart. Dash line (-), no tests conducted. Numerator is number of positive tests. Denominator is total number of tests conducted. On August 28, one individual returned to the dorm after completing isolation protocols.

Over the next two days, antigen tests (34 individuals) and PCR tests (24 individuals) were conducted for individuals not tested on August 26. All tests were negative (Table 2). Corresponding wastewater samples from August 27 and 28 were also negative (Table 1), indicating that the source(s) for SARS-CoV-2 had likely been removed from the dorm. However, wastewater analysis on August 29 was positive for both N1 (1.04 x 104 copies/L) and N2 (9.93 x 105 copies/L) genes (Table 1). From the TATS samples conducted on August 26, PCR results were positive for two tests three days later (Table 2), one of which was collected from Person A, who had previously tested positive via an antigen test and was already isolated. The other was a PCR retest that identified a new positive individual (Person C) whose earlier antigen test was negative and PCR was inconclusive. Person C was immediately relocated into isolation despite being asymptomatic.

Interestingly, SARS-CoV-2 RNA was not detected in wastewater on August 27 and 28 while Person C was in the dorm. Reports have indicated that approximately 50% of COVID-19 patients shed virus in feces. Therefore, it is possible that Person C was not shedding virus or was recently exposed and had low viral shedding on August 27 and 28. The low viral shedding load justification is supported by the fact that 40 cycles of PCR (C_q_ = 40) were required for the positive PCR result, which suggested trace amounts of SARS-CoV-2 *(8)*. Recent studies have also observed that viral loads from stool samples follow a more erratic pattern than viral loads from upper respiratory tract *(9,10)*. It is important to note that on August 28, Person D returned to the dorm after being on isolation protocols. Reports suggest that low viral shedding in feces can continue for over two weeks after symptoms have cease *(9,11–13)*. On August 29, viral RNA was detected in wastewater from Dorm A. This positive wastewater result is likely due to combined shedding from Person C and Person D. The removal of Person C on August 29 resulted in lower shedding loads, and ultimately, negative wastewater samples in the following days. Due to negative wastewater samples on August 30 and 31 (Figure 1; Table 1), no further clinical testing was performed.

## Conclusion

Overall, WBE combined with clinical testing successfully identified and potentially prevented COVID-19 transmission. Positive detection in wastewater samples always corresponded with positive clinical tests throughout the 12-day study. Thus, there were no false results, and positive detection in wastewater accurately signaled the presence of infected individuals in Dorm A. Infected individuals were identified via clinical tests enabling the university to initiate transmission intervention strategies (Figure 2). In contrast, WBE can result in a false negative result if an infected individual sheds little or no virus or if the environmental matrix results in PCR inhibition. Overall, this case study validates the utility of WBE to avert potential COVID-19 outbreaks.

**Figure 2.**
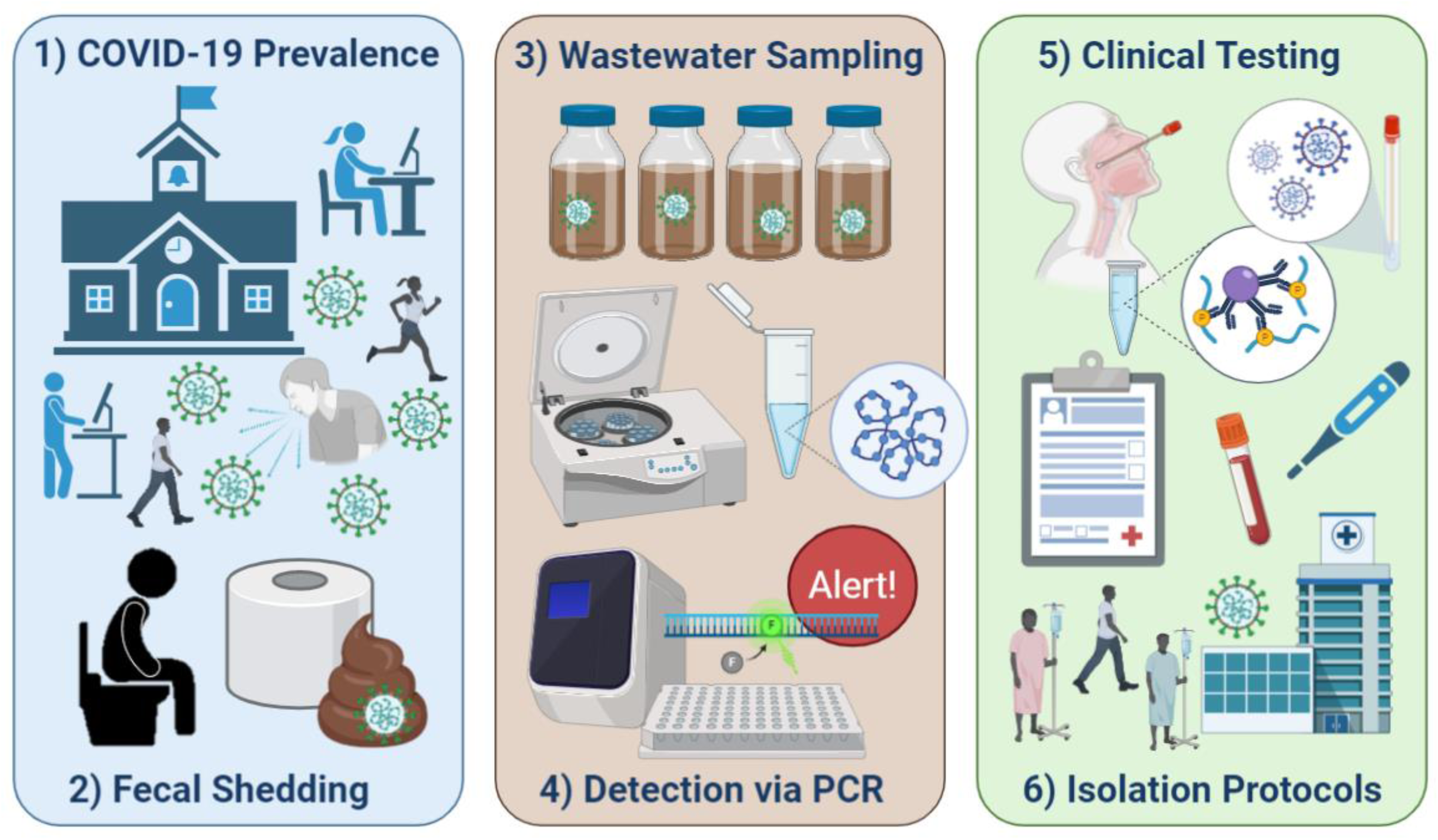
UArizona WBE and clinical testing protocol. Created with BioRender.com

## Data Availability

All data is available upon request to the corresponding author.

## Acknowledgments

The authors thank the UArizona Task Force, Amy Glicken, and Jeff Bliznick for their contributions. The work was supported by the University of Arizona Campus Re-Entry Plan. All relevant ethical guidelines have been followed and IRB approvals have been obtained.

## Author Bio

Walter Betancourt is an Environmental Virologist and Assistant Research Professor at The University of Arizona. His research is focused on detection and identification of waterborne viral pathogens and viral surrogates in water reuse systems.

## Appendix

### Design and Methods

#### Wastewater sampling

Wastewater samples were collected from a sewer manhole specific to Dorm A, without convergence or mixing from other sewer lines. Grab samples (1L) were collected from the manhole using a pole/dipper and submerging a sterile Nalgene bottle into the flowing wastewater until it was full. Samples were collected at 8:00 am on August 18 and 20 during the week that students moved into the dorm. Daily samples were collected from August 25 – 31 to monitor SARS-CoV-2 RNA in wastewater during the first week of classes. On August 26, five samples were collected five minutes apart between 8:30-8:50 am to determine sample variation during sample collection. On August 27, two samples were collected, on in the morning (9:30 am) and afternoon (1:15 pm) to determine variation at different times of day. All samples were transported in a cooler containing ice to the laboratory for immediate processing.

#### Virus concentration and recovery

The method for recovery and concentration of SARS-CoV-2 from wastewater was validated and standardized using human coronavirus 229E (HCoV 229E, ATCC VR-740) as a surrogate. Briefly, the initial virus concentration involved stepwise vacuum filtration through membrane filters of 0.8, 0.65, 0.45 and 0.22 µm pore sizes (EMD Millipore, Billerica, MA) followed by centrifugal ultrafiltration using the CentriconPlus-70 filter, 100 kDa cutoff (EMD Millipore, Billerica, MA). The final concentrate sample volume was used for RNA extraction as described below.

Matrix spikes were used to evaluate the performance and recovery efficiency of the method for concentration of enveloped viruses from wastewater samples from UArizona dormitories. The surrogate HCoV 229 E was spiked in dormitory sewage at concentrations of 1.30 x 10^10^ ± 2.97 x 10^9^ total gene copies per volume of sample. Aliquots of 0.5 L of raw sewage were spiked and processed following the method described above. Recovery efficiencies (Y) of HCoV 229 E were calculated as follows:

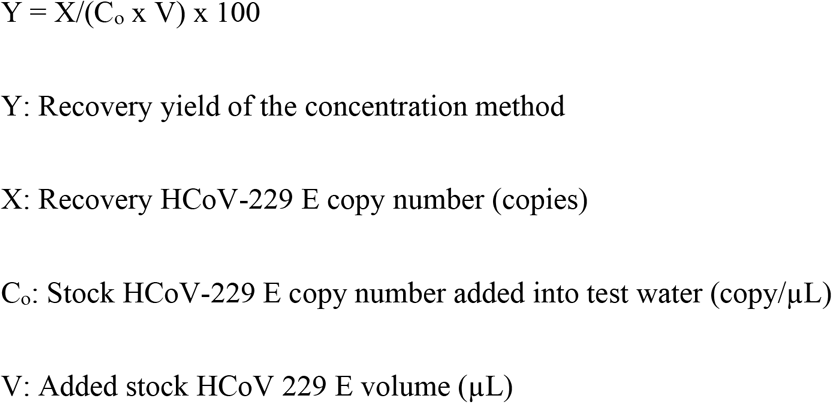

It is important to note that matrix spikes were included only in a baseline study in order to evaluate the recovery efficiency of the method. The concentrations of SARS-CoV-2 RNA were not adjusted to the estimated recovery efficiencies. These assays yielded an average recovery of 14±16% which indicated low and highly variable efficiencies of recovery of HCoV 229 E in wastewater as observed in studies with other coronaviruses used for the same purpose (*1-2*). Low recoveries of enveloped and non-enveloped viruses in wastewater and variations in the efficiency of the methods are predominantly associated with the complexity of this environmental matrix (*3-4*).

#### Virus Detection and Quantification

The nucleic acid of both SARS-CoV-2 and HCoV 229 E was extracted from the final concentrate sample volume using the QIAGEN QIAmp Viral Mini Kit (Qiagen, Valencia, CA) followed by cDNA synthesis from the extracted RNA using the SuperScript® IV First-Strand Synthesis reverse transcription kit (Invitrogen, Carlsbad, CA) with random hexamers. Samples were assayed for SARS-CoV-2 using the RT-PCR assays manufactured at Integrated DNA Technologies (IDT, Coralville, IA) for research use only (RUO). ROU kits include all published assays for the nucleocapsid genes N1 and N2 developed by the Centers for Disease Control and Prevention (Table S1). Similarly, samples were assayed for HCoV 229 E using a real-time quantitative reverse transcriptase PCR (RT-qPCR) assay previously developed for the rapid detection and quantitation of this virus in clinical samples (*5*).

Real-Time RT-PCR assays were performed using the LightCycler® 480 Instrument II (Roche Diagnostics, Indianapolis, IN). For SARS-CoV-2, reaction mixtures (25 µL) contained

12.5 µL of LightCycler 480 Probes Master (Roche Diagnostics) primers and probes (1.87 µL), nuclease-free water (5.63 µL) plus 5 µL of viral cDNA. For HCoV 229 E, reaction mixtures (25 µL) contained 12.5 µL of LightCycler 480 Probes Master (Roche Diagnostics) primers and probes (2.5 µL), nuclease-free water (5 µL) plus 5 µL of viral cDNA. Fluorescence data were collected after every cycle and analyzed with LightCycler® 480 Software version 1.5.1.6.2 (Roche Diagnostics). Primers and probes used for detection and quantitative estimation of SARS-CoV-2 RNA and HCoV 229E RNA in wastewater sample concentrates are described in Table S1.

For absolute quantification of SARS-CoV-2 and HCoV-229 E, standard curves were generated using ten-fold dilutions of a plasmid control containing the complete nucleocapsid gene from 2019-nCoV (IDT, Coralville, IA) and a gBlock gene fragment (IDT, Coralville, IA), respectively following the Roche system based on second-derivative C_q_determination and nonlinear fit algorithms. Limits of blank, detection, and quantification for the RT-qPCR assays were determined following standard procedures previously described (*6*) and currently in use in our laboratory. Limit of blank (LoB) are defined as the lowest concentration that can be reported with 95 percent confidence to be above the concentrations of blanks. The LoB was used to determine the most accurate limit of detection. It is the highest apparent concentration expected to be found when replicates of a blank are tested and is determined by calculating the 95th percentile of the C_q_values for all the blanks (reagent water containing no target material) for a specific target (C_q_LoB). This includes the C_q_ values for no-template controls, extraction blanks, and filter blanks. Limit of detection (LoD) is defined as the lowest concentration that can be detected with 95 percent confidence that it is a true detection and can be distinguished from the LoB. The LoD was determined by running a series of dilutions of the target with a minimum of 10 replicates per dilution. The dilution with the lowest concentration of known target that met the following requirements was chosen as the LoD: 1) the standard deviation (in Ct values) of the replicates was less than one and 2) the number of replicates with detections was greater than 95 percent. The average C_q_ value (C_q_LoD) for this dilution was used to calculate a concentration (copies/rxn) using the standard curve run with the dilution series. Limit of quantification (LoQ) is defined as the lowest concentration that can be accurately quantified. The LoQ was determined using the C_q_LoD and the standard deviation of C_q_LoD as previously defined. A C_q_ value for the LoQ (C_q_LoQ) was calculated as *C*_q_*LoQ* = *C*_q_*LoD* − 2(σ*C*_q_*LoD*) where σ*C*q*LoD* is the standard deviation of the C_q_LoD for this assay. This C_q_LoQ was used to calculate a concentration (copies/rxn) using the standard curve run with the dilution series. A summary of the performance of the standard curves for each assay is given in Table S2.

#### Clinical Testing and Public Health Protocols

UArizona performs two clinical tests for COVID-19 diagnosis: antigen (Sofia SARS Antigen FIA, Quidel, San Diego, CA, USA) via anterior nasal swab and RT-PCR (CDC 2019-nCoV RT-PCR Diagnostic Panel) (*7*) via nasopharyngeal swab samples. This test is not yet approved or cleared by the United States Food and Drug Administration. This test was developed and its analytical performance characteristics have been determined by the University of Arizona Genetics Core for Clinical Services. It has not been cleared or approved by FDA. This assay was developed as a Laboratory-Developed Test and has been validated pursuant to the CLIA regulations and is used for clinical purposes.

Upon arriving on campus, every individual was required to report to a designated COVID-19 testing site and undergo an anterior nasal swab for antigen testing. Individuals were kept on-site until tests results were noted. Each individual was required to test negative before receiving access to the dorm and campus buildings. If a person had a positive COVID-19 test, they were required to isolate for a minimum of 10 days (at home or in a designated isolation dorm) from the onset of symptoms or from the date the sample was taken, per guidelines from the United States Center for Disease Control and Prevention (*8*). If the individual remains symptom-free after 10 days, they can be cleared to return to the dorm; however, if symptoms persist they may be kept in isolation longer.

While living in the dorm, individuals were subject to clinical testing via two routes: Campus Health Services (CHS) or Test All Test Smart (TATS). CHS testing was conducted only on individuals that were experiencing symptoms and reported to the health services building for clinical testing and diagnosis. TATS testing was conducted on every individual living in the dorm upon positive detection of SARS-CoV-2 in wastewater that suggested prevalence of disease/infection amongst persons in the dorm. Individuals were excluded from TATS testing if they had already reported to CHS for testing on the same day or had proof of recently returning from isolation and no longer experiencing symptoms. In essence, CHS tested individuals that were symptomatic, and TATS tested individuals that were asymptomatic or had not yet reported symptoms. CHS and TATS both utilized anterior nasal swab samples for antigen and/or PCR tests.

**Table S1.**
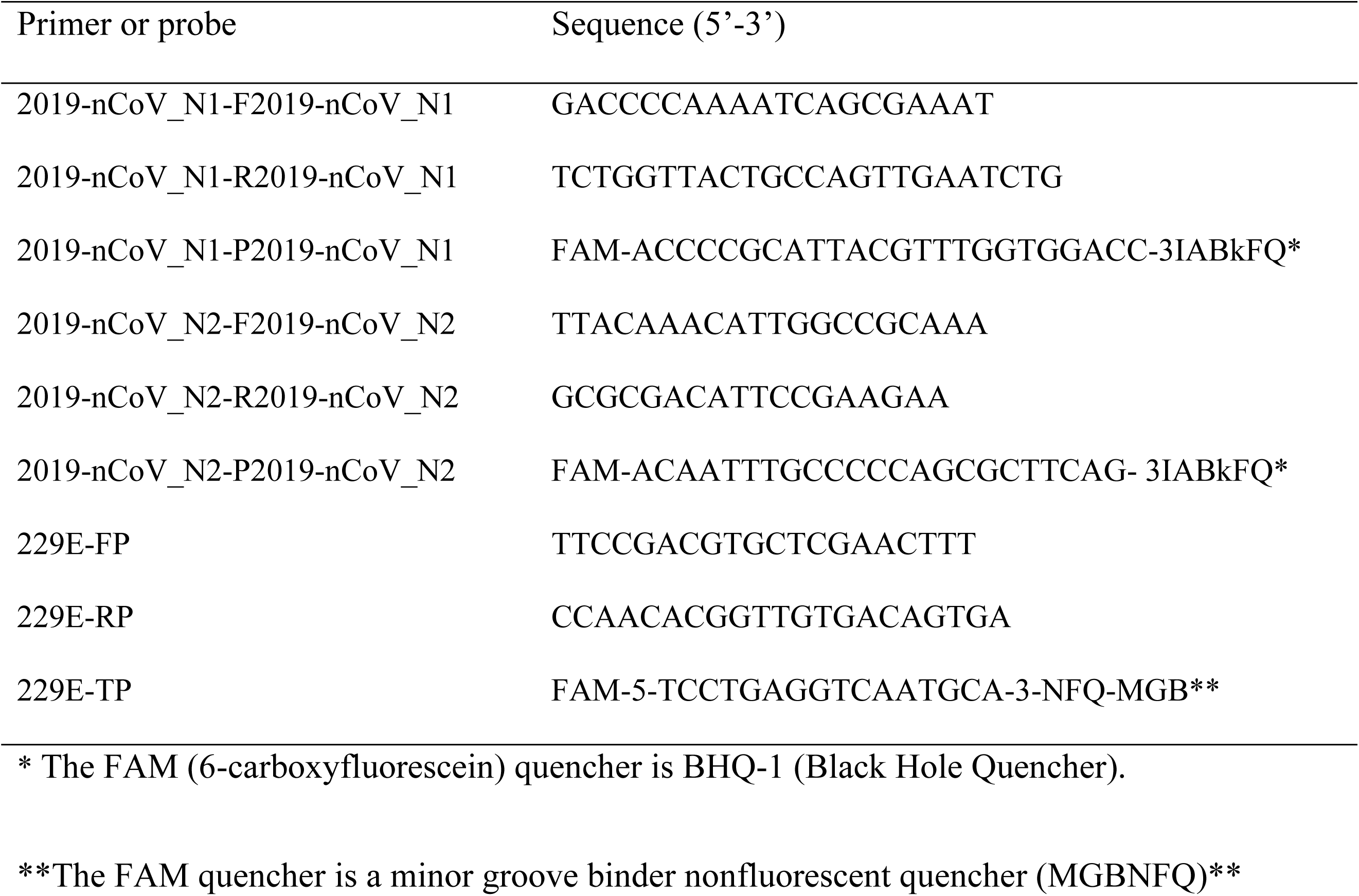
Primer and probe nucleotide sequences for SARS-CoV-2 and HCoV 229E

**Table S2.** Performance characteristics of the RT-qPCR standard curves

| Assay | Estimated LoQ* | Cq | Slope | y-intercept | Dynamic range<br>(copies/qPCR rxn) | Efficiency |
| --- | --- | --- | --- | --- | --- | --- |
| N1 | 1.30E+02 | (35.25 | -3.437 | 45.52 | 20 - 2000000 | 0.95 |
| N2 | 1.01E+04 | 35.51 | -4.482 | 53.46 | 2000 - 2000000 | 0.65 |
| 229E | 3.86E+02 | 36.85 | -3.470 | 45.80 | 250 - 2500000 | 0.95 |

\* LoQ = limit of quantification

**Table S3.** CT values for LoD positive control sample relative to accompanying nCoVPC serial diltion of known concentration.

| Instrument | Assay | nCoVPC serial dilution - viral copies/reaction |  |  |  |  |  |  |  |  |  | Ave | sd |
| --- | --- | --- | --- | --- | --- | --- | --- | --- | --- | --- | --- | --- | --- |
|  |  | 10000 | 5000 | 2500 | 1250 | 625* | 313* | 156 | 78 | 39 | 20 |  |  |
| LC480 | N1 | 28.26 | 29.28 | 30.52 | 31.49 | 32.46 | 33.76 | 35.78** | 36.57 | 35.78 | 37.46 | 33.63 | 0.44 |
|  | N2 | 31.05 | 32.36 | 33.99 | 35.83 | 37.30 | 38.44 | 40.00** | 40.00 | 40.00 | - | 37.29 | 0.72 |
| 7900 | N1 | 27.89 | 28.63 | 29.96 | 30.73 | 32.48 | 33.51 | 34.59 | 37.38 | 37.71 | 35.58 | 33.21 | 0.50 |
|  | N2 | 29.00 | 29.80 | 30.33 | 31.96 | 33.75 | 34.34 | 34.88 | 35.03 | 40.29 | - | 34.20 | 0.56 |

\* Limit of detection (LoD) average CT fall between Ct values of nCoVPC serial dilution control.
\*\* Range of CTs shown in patient sample.

